# Factors Influencing Road Safety Protocol Compliance among Commercial Vehicle Drivers at Hohoe in the Volta Region of Ghana

**DOI:** 10.1101/2025.09.04.25335130

**Authors:** Abigail Asabea Addo, Francis Arizie, Anthony Edward Boakye, Veronica Okwuchi Charles-Unadike

**Affiliations:** Department of Biostatistics and Epidemiology, University of Health and Allied Sciences, Ho, Ghana., Contribution: Conceptualize the study, Methodology, Formal analysis, Data curation, Writing – original draft; Department: Research office, Business and Health, York St John University., Contribution: Formal analysis, Data curation, Writing – original draft, and Resources; Department of Health, Physical Education and Recreation, University of Cape Coast, Cape Coast, Ghana, Contribution: Methodology, Formal analysis, Writing – original draft, and Writing – review & editing; Department of Population and Behavioural Science, School of Public Health, University of Health and Allied Sciences, Ho Volta Region, Ghana., Contribution: Methodology, Supervision, and writing review and editing

**Keywords:** Adherence, Passenger drivers, Predictors, Road, Safety measures, Vehicle

## Abstract

**Background:** The prevalence of compliance with road safety regulations among commercial motorcycle riders at Hohoe is observed to be moderate with a rate of 59.2%. This implies that while a significant proportion of riders adhere to some safety guidelines, others still engage in practices that could increase their risk of accidents.

**Objective:** In line with this, the study aimed to investigate the determinants of road safety protocol compliance among commercial vehicle drivers at Hohoe in the Volta Region of Ghana.

**Methods:** The study was structured in a positivist worldview and adopted census method with cross-sectional descriptive design. Data were solicited from 395 participants from the field and were processed with SPSS version 27. Frequency distribution, chi-square test, and binary logistic regression were used to analyse the data. The frequency distribution was used to summarise participants responses into proportions. The Pearson’s chi-square test of independence was used to test the hypotheses postulated in the study, either to accept or reject the null hypotheses. However, the binary logistic regression was used to determine the effects of the IVs on the DV.

**Results:** The study found that Islamic religion was less significantly related to road safety protocol compliance. It emerged that associations were found between socio-demographic characteristics (*p*<0.001), human factors (*p*<0.001), vehicle factors (*p*<0.001), environmental factors (*p*<0.001) as well as road factors (*p*<0.001), and commercial drivers’ road safety protocol compliance at Hohoe in the Volta Region of Ghana.

**Conclusion:** These findings suggest that improving compliance requires a multifaceted approach that prioritises infrastructure development, driver training, and targeted interventions based on the broader contextual factors impacting driver behaviour, rather than focusing solely on religious affiliation.

## Introduction

Road traffic deaths and injuries remain a major global health and development challenge [1–3]. Globally, there were an estimated 1.19 million road traffic deaths in 2021 – a 5% decrease when compared with the 1.25 million deaths in 2010 [1–3]. As of 2019, road traffic crashes were noted to be the leading cause of death of children and youth age 5 to 29 years, and were claimed to be the 12th leading cause of death when all ages are considered [1,2,4]. Two-thirds of deaths occur among people of working age (18–59 years), causing huge health, social and economic harm throughout society [1,5]. It was established that 92% of these deaths occur in low- and middle-income countries [1,6,7]. The risk of death is three times higher in low-income countries than high-income countries despite these countries having less than 1% of all motor vehicles [1,8,9].

In 2023, Ghana recorded 14,135 road crashes, resulting in 2,276 fatalities and 15,409 injuries [10]. Therefore, to reduce accident risks, and foster public trust, maintaining road safety compliance is essential. Compliance refers to drivers adhering to traffic rules as well as regulations and laws to ensure safe and responsible driving [11]. However, to stay on top of compliance, it is essential for commercial vehicle operators to understand, and implement specific regulations, maintaining vehicle standards, and regularly monitoring both driver behaviour and vehicle conditions [11]. Evidence suggests that at Hohoe, within the first half of the 2022, only 7 crashes were recorded representing 3.6% of the cases within the Volta Region of Ghana of which no fatality was witnessed [12]. The prevalence of compliance with road safety regulations among commercial motorcycle riders at Hohoe is observed to be moderate with a rate of 59.2% [13]. This implies that while a significant proportion of riders adhere to some safety guidelines, others still engage in practices that could increase their risk of accidents [13].

In pursuit of identifying studies that are peer reviewed and published in reputable journals on the phenomena understudy, revealed that there are extant literatures in the field. However, it will interest you to note that those studies focused predominantly on factors contributing to road crashes among commercial vehicle drivers [14–19] leaving a knowledge gap on the determinants of road safety compliance among commercial vehicle drivers. In line with this, the study aimed to investigate the determinants of road safety protocol compliance among commercial vehicle drivers at Hohoe in the Volta Region of Ghana.

### Specifically, the study seeks to

1. Examine if socio-demographic factors of commercial vehicle drivers influence road safety protocol compliance at Hohoe in the Volta Region of Ghana;
2. Ascertain the extent to which human factors influence road safety protocol compliance among commercial vehicle drivers at Hohoe in the Volta Region of Ghana;
3. Analyse if vehicle factors influence road safety protocol compliance among commercial vehicle drivers at Hohoe in the Volta Region of Ghana;
4. Assess whether environmental factors predict road safety protocol compliance among commercial vehicle drivers at Hohoe in the Volta Region of Ghana; and lastly,
5. Determine whether road factors influence road safety protocol compliance among commercial vehicle drivers at Hohoe in the Volta Region of Ghana.

The study further hypothesised that there is no statistically significant relationship between socio-demographic, human, vehicle, environmental as well as road factors, and road safety protocol compliance among commercial vehicle drivers at Hohoe in the Volta Region of Ghana.

## Methods

### Study philosophy and design

The study was structured in a positivist worldview and lends itself to a quantitative approach. This worldview was preferred because it relies on measurement and reason, that knowledge is revealed from a neutral and measurable (quantifiable) observation of activity, action or reaction [20,21]. With this approach, a descriptive cross-sectional study design was deemed appropriate. The design was suitable due to the relatively low cost involved and its ability to collect data within a relatively short time and at a single point period [22,23]. It also offers the opportunity to simultaneously measure exposure and outcomes [24]. Nevertheless, it has some disadvantages which include its inability to determine cause and effect, also it cannot be used to assess a specific behaviour over a period [24].

### Study setting and population

The study was conducted between September 2022 and ended on September 2023 at Hohoe (a municipal capital) in the Volta Region of Ghana. Hohoe has four transport unions - Ghana Private Road Transport Union (GPRTU), Global Millennium Transport Association (GMTA), Co-operative Transport Union (CO-OP), and Progressive Transport Union (PROTOA) [25]. The main means of transportation for the inhabitants of Hohoe is by commercial vehicles, which are taxis, for the inner cities and mini busses for journeys outside Hohoe. However, in the year 2022, Hohoe, Kpando, and Ketu divisions recorded 7 cases each in the first half of the year which was the least. Further, with this being the least, Hohoe division did not record any fatality in that same period [26]. These statistics ignited the study to investigate the factors that contributed to road safety protocol compliance among commercial drivers to achieve that excellent record in the first half of 2022. The population for the study was all commercial drivers age 20 years and above who have their vehicles registered for commercial use only.

### Data source and sampling technique

Data for the study were solicited from 395 participants from the field with questionnaire. The questionnaire was developed based on literature and standardised instruments used by other researchers in previous studies. Further, the questionnaire was divided into five sections. Section 1 occupies socio-demographic characteristics of the participants; section 2 covered items on human factors; section 3 contains vehicle factors; section 4 occupies items on environmental factors; while the last section covered items on road factors. The study employed census sampling method which permitted us to collect data from all the members of the population of interest. Moreover, it is accurate, reliable and help researchers to eliminate sampling error [27].

### Measures

In this study, the Independent Variables (IVs) are socio-demographic characteristics, human factors, vehicle factors, environmental factors, and road factors while the Dependent Variable (DV) is road safety protocol compliance. The IVs were carefully chosen because we aimed to investigate the influence they have on commercial drivers’ road safety protocol compliance. Hence, previous studies have not examined the interplay of the variables together [28–32].

### Reliability and validity

Effort was made to ensure reliability and validity in the study. On reliability, the data collection instruments were pretested on 50 commercial drivers from Oti Region just to ensure that they were devoid of ambiguity and also measure what they opt to measure. However, Cronbach alpha reliability test was also conducted on the data to ascertain its acceptability for analysis. Cronbach alpha coefficient of 0.60 was obtained which was an indication that the data were acceptable for analysis. This was supported by the general rule of thumb that a Cronbach’s alpha of 0.60-0.70 is acceptable and that data is good for analysis [33].

For face validity, the questionnaire was given to colleague students to read through to help correct grammatical errors and also check to see if the items were structured in accordance with the objectives. The feedback obtained helped in restructuring the questionnaire. Further, the questionnaire was given to expert in the field (environmental health and occupational safety) to also ascertain if they were appropriate to measure the objective of the study. Aside that, the processes such as designing of questionnaire, choosing of research methods, statistical tools for analysis and so forth were keenly taken care of since we did not want these processes to affect the findings of the study [34].

### Data Collection Procedure

Although, census method was used for the study. However, since this study was not a traditional government census study which counts every person in a population, it became essential for participants to be recruited. Therefore, recruitment of the study participants commenced on Monday, 10th of October, 2022 and ended on 31^st^ of January, 2023 after The Research Committee of the University of Health and Allied Sciences had reviewed the research protocol and unanimously granted an ethical clearance with ID UHAS-REC A. [018] 21-22 on 28^th^ of September, 2022 to carry out the study. After the recruitment of participants, the actual fieldwork took place on Friday 10^th^ of February, 2023 and ended on Saturday 25^th^ of February, 2023. In all, two weeks were used to collect the data from the field. Four research assistants were hired and trained to assist in the data collection. In the field, research assistants were assisted with computer tablets to collect the data.

### Data processing and analysis

Data collected from respondents were cleaned and edited. Items that requested multiple responses were recoded for easy entry. SPSS version 27 was used to process the data. Frequency distribution, chi-square test, and binary logistic regression were used to analyse the data. The frequency distribution was used to summarise participants’ responses into proportions. The Pearson’s chi-square test was used to test the hypotheses postulated in the study, either to accept or reject the null hypotheses. The binary logistic regression was used to determine the factors associated with road safety protocol compliance among commercial vehicle drivers.

### Ethical Considerations

The study tried to ensure that it meets ethical standards. In view of this, the study protocol was submitted to The Research Committee of the University of Health and Allied Sciences for review and the university unanimously approved the research protocol and granted an ethical clearance with ID (UHAS-REC A. [018]) 21-22. However, in the field, confidentiality, anonymity, and privacy were ensured. On confidentiality, participants were informed that the information they provided would be kept confidential from third parties. The information given is strictly for academic purposes and that the findings would be published to add to academic literature in the field of environmental health and occupational safety. On anonymity, we tried to avoid anything that could tag a participant to a questionnaire which would invariably leads his information to other people such as name, address, and telephone numbers while on privacy, participants were made to choose a place they deem appropriate for the structured interview. Further, participation was made voluntary and participants were briefed that they reserve the right to withdraw from the study if they wish to do so without any consequences. Furthermore, before a participant takes part in the study, an oral (verbal) informed consent was taken. Though we did not document the verbal consent given by the participants but it was witnessed by their various association leaders including the chairman. As part of the inclusion criteria was age limit of 20 years hence, no minor participated in the study.

## Results

The participants sampled, more than twenty percent (23.3%) were between 50 -to-59 years age group category while 16.5% were either between 20-to-29 or 40-to-49 years age group. Whereas Christianity was the dominant category constituting 61.8%, the least category was traditional (11.1%). About third (33.2%) belong to five and above member household while 16.5% belong to 2-member household. More than half (55.4%) have been in operation for more than six years while 5.8% have been in operation for less than a year (See Table 1).

**Table 1:** Number of Drivers in the Various Transport Unions at Hohoe.

| Transport unions | Number of Members |
| --- | --- |
| GPRTU | 250 |
| PROTOA | 36 |
| CO-OP | 58 |
| GMTA | 51 |
| **Total** | **395** |

Source: Fieldwork (2023).

**Table 1:**
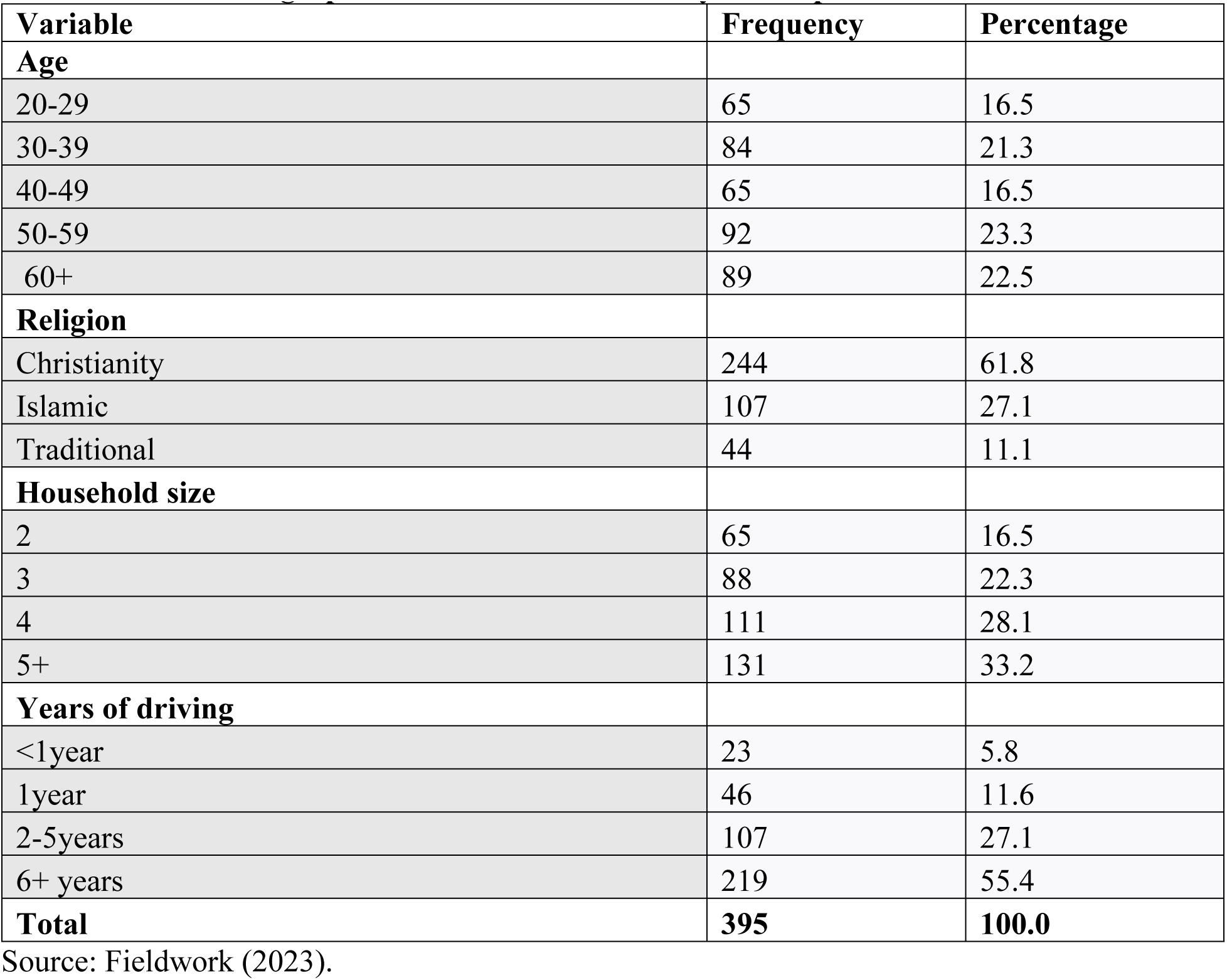
Socio-Demographic Characteristics of Study Participants.

To identify the number of drivers who comply with road safety protocol instigated us to ask a global yes/no question thus (always comply with road safety protocol) which is used to ascertain commercial drivers who do comply with road safety protocol. After analysis, the results revealed that 351(89%) of the commercial vehicle drivers answered in the affirmative while 44(11%) of them intimated they do not comply with road safety protocol.

Table 2 presents Pearson’s chi-square test of independence results on socio-demographic characteristics and commercial drivers’ road safety protocol compliance at Hohoe in the Volta Region of Ghana. This analysis was conducted to test the hypothesis there is no statistically significant relationship between socio-demographic characteristics and commercial drivers’ road safety protocol compliance at Hohoe in the Volta Region of Ghana. Statistically significant relationships were found between age [𝛘^2^=58.634, *p*<0.001], religion [𝛘^2^=14.025, *p*<0.001], household [𝛘^2^=79.757, *p*<0.001] as well as years of driving [𝛘^2^=39.793, *p*<0.001] and commercial drivers’ road safety protocol compliance at Hohoe in the Volta Region of Ghana.

**Table 2:** Relationship between Socio-Demographic Characteristics and Commercial Drivers’ Road Safety Protocol Compliance at Hohoe in the Volta Region of Ghana.

| Variable | No (%) | Yes (%) | Total n (%) | $\chi^2$ | P-value |
| --- | --- | --- | --- | --- | --- |
| **Age** |  |  |  | 58.634 | <0.001 |
| 20-29 | 0.0 | 100.0 | 65(100.0) |  |  |
| 30-39 | 0.0 | 100.0 | 84(100.0) |  |  |
| 40-49 | 0.0 | 100.0 | 65(100.0) |  |  |
| 50-59 | 25.0 | 75.0 | 92(100.0) |  |  |
| 60+ | 23.6 | 76.4 | 89(100.0) |  |  |
| **Religion** |  |  |  | 14.025 | <0.001 |
| Christianity | 9.4 | 90.6 | 244(100.0) |  |  |
| Islamic | 19.6 | 80.4 | 107(100.0) |  |  |
| Traditional | 0.0 | 100.0 | 44(100.0) |  |  |
| **Household size** |  |  |  | 79.757 | <0.001 |
| 2 | 32.3 | 67.7 | 65(100.0) |  |  |
| 3 | 26.1 | 73.9 | 88(100.0) |  |  |
| 4 | 0.0 | 100.0 | 111(100.0) |  |  |
| 5+ | 0.0 | 100.0 | 131(100.0) |  |  |
| **Years of driving** |  |  |  | 39.793 | 0.001 |
| <1year | 0.0 | 100.0 | 23(100.0) |  |  |
| 1year | 0.0 | 100.0 | 46(100.0) |  |  |
| 2-5years | 0.0 | 100.0 | 107(100.0) |  |  |
| 6+ years | 20.1 | 79.9 | 219(100.0) |  |  |

Note: Row percentages in parenthesis, Chi-square significant at (0.001), (0.05), (0.10)

Further analysis was conducted with binary logistic regression on socio-demographic characteristics and commercial drivers’ road safety protocol compliance at Hohoe in the Volta Region of Ghana. This analysis was conducted to ascertain the influence of socio-demographic characteristics on commercial drivers’ road safety protocol compliance at Hohoe in the Volta Region of Ghana (See Table 3).

**Table 3:** Binary Logistic Regression on Socio-Demographic Characteristics and Commercial Drivers’ Road Safety Protocol Compliance at Hohoe in the Volta Region of Ghana.

| Variable | B | Wald | Sig. | Exp(B) | 95CI |  |
| --- | --- | --- | --- | --- | --- | --- |
| **Religion** (Christianity=1.0) |  |  |  |  |  |  |
| Islamic | -0.853 | 6.782 | 0.009 | 0.426 | 0.224 | 0.810 |
| Traditional | 18.940 | 0.000 | 0.998 | 168126341.111 | 0.000 | 0.000 |
| **Constant** | **2.263** | **106.653** | **0.000** | **1. 9.609** | 2. | 3. |

4. Source: Fieldwork (2023). Significant at 0.05.

After processing the data, only one variable (religion) was significant. Those that were not significant were removed from the model (see Table 3). Overall, the logistic regression model was significant at −2LogL = 258.365; Nagelkerke R^2^ of 0.87; 𝝌^2^= 17.674; *p*<0.001 with correct prediction rate of 88.9%. More importantly, the Model Summary which shows a Nagelkerke R^2^ of 0.87 suggests that the model explains 87% of variance in the likelihood of commercial drivers’ road safety protocol compliance at Hohoe in the Volta Region of Ghana. With this percentage contribution to the entire model, the results confirmed the whole model significantly predict commercial drivers’ road safety protocol compliance at Hohoe in the Volta Region of Ghana. Table 3 revealed that Islamic was significantly related to road safety protocol compliance at *p*<0.001, (OR=0.426, 95%CI ([0.224-0.810]). This variable identifies those commercial drivers to have 0.43 times less likely to comply with road safety protocol compared with their Christian counterparts (see Table 3). However, traditional was not significant which is an indication that road safety protocol compliance among commercial drivers at Hohoe in the Volta Region of Ghana is not predicted by one’s affiliation with the traditional religion.

To find responses for research objective 2 which is “what is the extent to which human factors influence road safety protocol compliance among commercial vehicle drivers at Hohoe in the Volta Region of Ghana” triggered us to ask a question revolving human factor leading to road safety protocol compliance. After analysis, the results revealed that 33% of the participants reported that it is physical and mental conditions while 17.0% indicated that it is drivers’ skills and experience (See Table 4).

**Table 4:** Human Factors leading to Road Safety Compliance.

| Variable | Frequency | Percentage |
| --- | --- | --- |
| **Human factors** |  |  |
| Drivers behavior | 111 | 28.1 |
| Drivers skills and experience | 67 | 17.0 |
| Psychological factors | 88 | 22.3 |
| Physical and mental conditions | 129 | 32.7 |
| **Total** | **395** | **100.0** |

Source: Fieldwork (2023).

In Table 5, has the outcome of Pearson’s chi-square test of independence on human factors and commercial vehicle drivers’ road safety protocol compliance at Hohoe in the Volta Region of Ghana. This analysis was essential to test the hypothesis there is no statistically significant relationship between human factors and commercial vehicle drivers’ road safety protocol compliance at Hohoe in the Volta Region of Ghana. Statistically significant relationship was found between human factors [𝝌^2^=33.168, *p*<0.001] and commercial vehicle drivers’ road safety protocol compliance at Hohoe in the Volta Region of Ghana.

**Table 5:**
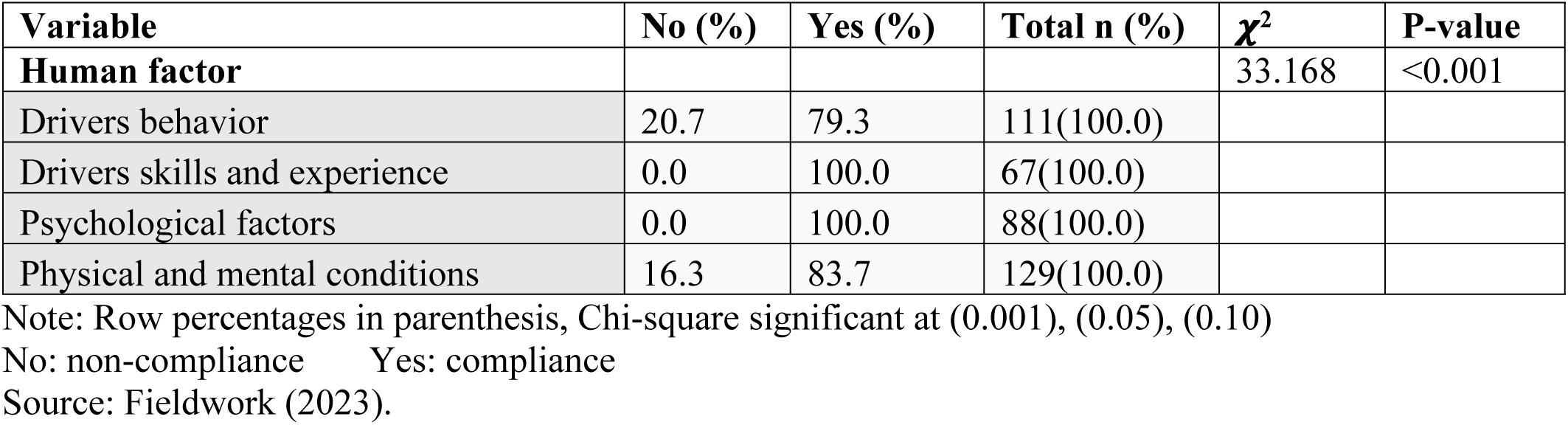
Relationship between Human Factors and Commercial Vehicle Drivers’ Road Safety Protocol Compliance at Hohoe in the Volta Region of Ghana.

| Variable | No (%) | Yes (%) | Total n (%) | $\chi^2$ | P-value |
| --- | --- | --- | --- | --- | --- |
| **Human factor** |  |  |  | 33.168 | <0.001 |
| Drivers behavior | 20.7 | 79.3 | 111(100.0) |  |  |
| Drivers skills and experience | 0.0 | 100.0 | 67(100.0) |  |  |
| Psychological factors | 0.0 | 100.0 | 88(100.0) |  |  |
| Physical and mental conditions | 16.3 | 83.7 | 129(100.0) |  |  |

Note: Row percentages in parenthesis, Chi-square significant at (0.001), (0.05), (0.10)
No: non-compliance Yes: compliance
Source: Fieldwork (2023).

To identify if vehicle factors trigger road safety protocol compliance among commercial vehicle drivers at Hohoe in the Volta Region of Ghana prompted us to ask a question revolving vehicle factor. The results found revealed that 39% of the participants reported vehicle conditions while 28% indicated tire condition (See Table 6).

**Table 6:** Vehicle Factors leading to Road Safety Protocol Compliance.

| Variable | Frequency | Percentage |
| --- | --- | --- |
| **Vehicle factors** |  |  |
| Vehicle conditions | 153 | 38.7 |
| Vehicle type | 131 | 33.2 |
| Tire Condition | 111 | 28.1 |
| **Total** | **395** | **100.0** |

Source: Fieldwork (2023).

Table 7 presents outcome of Pearson’s chi-square test of independence on vehicle factors and commercial vehicle drivers’ road safety protocol compliance at Hohoe in the Volta Region of Ghana. This analysis was significant hence, it was used to test the hypothesis there is no statistically significant relationship between vehicle factors and commercial vehicle drivers’ road safety protocol compliance at Hohoe in the Volta Region of Ghana. Statistically significant relationship was found between vehicle factors [𝝌^2^**=**32.641, *p*<0.001] and commercial vehicle drivers’ road safety protocol compliance at Hohoe in the Volta Region of Ghana.

**Table 7:** Relationship between Vehicle Factors and Commercial Vehicle Drivers’ Road Safety Protocol Compliance at Hohoe in the Volta Region of Ghana.

| Variable | No (%) | Yes (%) | Total n (%) | $\chi^2$ | P-value |
| --- | --- | --- | --- | --- | --- |
| **Vehicle factors** |  |  |  | 32.641 | <0.001 |
| Vehicle conditions | 0.0 | 100.0 | 153(100.0) |  |  |
| Vehicle type | 16.0 | 84.0 | 131(100.0) |  |  |
| Tire Condition | 20.7 | 79.3 | 111(100.0) |  |  |

Note: Row percentages in parenthesis, Chi-square significant at (0.001), (0.05), (0.10)
No: non-compliance Yes: compliance
Source: Fieldwork (2023).

To unravel environmental factors that lead to road safety protocol compliance among commercial vehicle drivers at Hohoe in the Volta Region of Ghana, made us to ask a question revolving environmental factor. The results obtained, revealed that about 45% of the participants reported it is road conditions while 22.3% indicated that it is lighting conditions (See Table 8).

**Table 8:** Environmental Factors leading to Road Safety Protocol Compliance.

| Variable | Frequency | Percentage |
| --- | --- | --- |
| **Environmental factors** |  |  |
| Road conditions | 176 | 44.6 |
| Time of day | 131 | 33.2 |
| Lighting conditions | 88 | 22.3 |
| **Total** | **395** | **100.0** |

Source: Fieldwork (2023).

In Table 9, has Pearson’s chi-square test results on environmental factors and commercial vehicle drivers’ road safety protocol compliance at Hohoe in the Volta Region of Ghana. This analysis was conducted to test the hypothesis there is no statistically significant relationship between environmental factors and commercial vehicle drivers’ road safety protocol compliance in the Hohoe Municipality of Ghana. Statistically significant relationship was found between environmental factors [𝝌^2^=99.787, *p*<0.001] and commercial vehicle drivers’ road safety protocol compliance at Hohoe in the Volta Region of Ghana.

**Table 9:** Relationship between Environmental Factors and Commercial Vehicle Drivers’ Road Safety Protocol Compliance at Hohoe in the Volta Region of Ghana.

| Variable | No (%) | Yes (%) | Total n (%) | $\chi^2$ | P-value |
| --- | --- | --- | --- | --- | --- |
| **Environmental factors** |  |  |  | 99.787 | <0.001 |
| Road conditions | 0.0 | 100.0 | 176(100.0) |  |  |
| Time of day | 33.6 | 66.4 | 131(100.0) |  |  |
| Lighting conditions | 0.0 | 100.0 | 88(100.0) |  |  |

Note: Row percentages in parenthesis, Chi-square significant at (0.001), (0.05), (0.10)
No: non-compliance Yes: compliance
Source: Fieldwork (2023).

In pursuit of ascertaining if road factors lead to road safety protocol compliance among commercial vehicle drivers at Hohoe in the Volta Region of Ghana, instigated us to ask a question revolving road factor. The results found revealed that about 39% of the participants reported traffic volume while about 28% indicated road design: sharp curves, narrow lanes, and/or inadequate signage (See Table 10).

**Table 10:** Road Factors leading to Road Safety Protocol Compliance.

| Variable | Frequency | Percentages |
| --- | --- | --- |
| **Road factors** |  |  |
| Road design: sharp curves, narrow lanes, or inadequate signage | 110 | 27.8 |
| Traffic volume | 153 | 38.7 |
| Road construction and maintenance | 132 | 33.4 |
| **Total** | **395** | **100.0** |

Source: Fieldwork (2023).

Table 11 presents the outcome of Pearson’s chi-square test of independence on road factors and commercial vehicle drivers’ road safety protocol compliance at Hohoe in the Volta Region of Ghana. This analysis was carried out to test the hypothesis there is no statistically significant relationship between road factors and commercial vehicle drivers’ road safety protocol compliance at Hohoe in the Volta Region of Ghana. Statistically significant relationship was found between road factors [𝝌^2^=20.091, *p*<0.001] and commercial vehicle drivers’ road safety protocol compliance at Hohoe in the Volta Region of Ghana.

**Table 11:** Relationship between Road Factors and Commercial Vehicle Drivers’ Road Safety Protocol Compliance at Hohoe in the Volta Region of Ghana.

| Variable | No (%) | Yes (%) | Total n (%) | $\chi^2$ | P-value |
| --- | --- | --- | --- | --- | --- |
| **Road factors** |  |  |  | 20.091 | <0.001 |
| Road design: sharp curves, narrow lanes, or inadequate signage | 0.0 | 100.0 | 110(100.0) |  |  |
| Traffic volume | 13.7 | 86.3 | 153(100.0) |  |  |
| Road construction and maintenance | 17.4 | 82.6 | 132(100.0) |  |  |

Note: Row percentages in parenthesis, Chi-square significant at (0.001), (0.05), (0.10)
No: non-compliance Yes: compliance
Source: Fieldwork (2023).

## Discussion

The study aimed to investigate the factors influencing road safety protocol compliance among commercial vehicle drivers at Hohoe in the Volta Region of Ghana. The findings reveal that Islamic religion tend to have lower odds of road safety protocol compliance. This finding was consistent with a previous study which found that the average daily traffic accidents (p-value = 0.040) and injuries (p-value = 0.001) increased significantly in the month of Ramadan [35]. This finding suggests that these drivers do not recognise the road safety protocol and that are not obliged to comply with them [36]. It could also suggest that the drivers are of the view that even if they comply with the road safety protocol, it cannot prevent them from road traffic accidents. The plausible explanation to this finding could be that participants affiliated to the Islamic religion despise speeding, do not engage in any behaviour such as eating, making phone calls, texting and so forth that would distract their attention while driving, and also adhere to car maintenance culture which invariably make them to overlook at the road safety protocols [37].

The study found that relationship exists between socio-demographic characteristics and commercial drivers’ road safety protocol compliance at Hohoe in the Volta Region of Ghana. Therefore, the null hypothesis was ignored. This finding corroborated with previous studies which found that demographic characteristics relate to attitudes towards traffic safety and pedestrian behaviours [38,39]. A p-value of <0.001 found in all the indicators suggests that both the socio-demographic characteristics and road safety protocol compliance are not independence of each other, and that they are interdependence. Further, the relationship found is an indication that socio-demographic characteristics predict road safety protocol compliance among commercial vehicle drivers at Hohoe in the Volta Region of Ghana [29].

It appeared relationship existed between human factors and commercial vehicle drivers’ road safety protocol compliance at Hohoe in the Volta Region of Ghana due to this, the null hypothesis was rejected. This outcome confirms previous studies which found that human factors such as speeding, aggressive driving and lastly fatigue were the main causes of road traffic accidents [40,41]. This relationship indicates that the variables studied are not independence of each other. Further, the relationship suggests that human factors influence drivers at Hohoe in the Volta Region of Ghana to comply with road safety protocol. The p-value (<0.001) found indicates a strong relationship between the variables tested [42].

It emerged that relationship exists between vehicle factors and commercial vehicle drivers’ road safety protocol compliance at Hohoe in the Volta Region of Ghana. Based on this, the null hypothesis was not confirmed. This finding is in line with previous studies, which found that many factors influence the road safety of which the most important factors are driver behaviour, construction and condition of the vehicle and condition of infrastructure [14,43]. A p-value of <0.001 found is an indication that the variables are not independence of each other hence, they interact together. The finding suggests that commercial vehicle drivers’ road safety protocol compliance is largely predicted by vehicle factors [43].

After testing environmental factors against commercial vehicle drivers’ road safety protocol compliance at Hohoe in the Volta Region of Ghana revealed that relationship existed between them, due to this, the null hypothesis was disproved. This finding agrees with previous studies, which found that weather conditions significantly impact road safety, with fog, rain, snow, and ice causing decreased visibility and increased risk [44,45]. The authors further stressed that floodwaters cause damage to road surfaces, leading to potholes and cracks. Snow/ice conditions make driving more treacherous, making skidding or losing control an imminent risk. Overall, weather conditions significantly influence road safety [44,45]. The relationship found indicates that both environmental factors and road safety protocol compliance are not independence of each other, and that they are interdependence. The finding suggests that road safety protocol compliance among commercial drivers is strongly predicted by environmental factors [17].

It emerged that road factors and commercial vehicle drivers’ road safety protocol compliance were revealed to be interrelated, due to this, the null hypothesis was not accepted. This outcome is in line with previous studies which found that poor road design, inadequate road maintenance, and confusing or missing traffic signs can lead to road accidents [17,43,46]. This finding suggests that road factors play a significant role in road safety protocol compliance among commercial drivers at Hohoe in the Volta Region of Ghana [15].

Per the findings of the study, 89% of the commercial vehicle drivers comply with road safety protocol. This finding corroborated with a previous study which found that overall, 5.3% of vehicles ran red lights—meaning nearly 95% of drivers stopped correctly, reflecting high compliance behaviour on signal-controlled roads. Compliance was slightly lower in Kumasi (about 93 %) and higher in Accra (approximately 95.6 %) [47]. This outcome suggests that overwhelming majority of commercial drivers are law abiding citizens which invariably transcended into them complying with road safety protocols [48,49]. Further, it could also suggest that they uphold their occupation in high esteem and that do not want to compromise with the safety of passengers [49]. It could also suggest that they normally get terrified upon hearing the sudden increase of road traffic accident-related fatalities, and injuries [16,50–52]. The plausible explanation for this finding could be that these commercial drivers want to reduce the number of motor vehicle crash they do record to minimal [16]. It could also be that the various associations they belong has educated them well on road safety protocols and why there is a need for them to comply with them [49,52].

However, the 11% who intimated they do not comply with road safety protocol reason could be that they have been in operation for long, and that thought they are experienced. Therefore, they do not need to comply with any road safety protocol before they can diligently carry out their duties [53]. This finding suggests that a significant proportion of the commercial drivers are not moved by the rising numbers of road traffic accident related injuries and casualties [28,54]. This outcome refuted a previous study which found that overall non-compliance to traffic rules was 25 - 30% [55].

It emerged that 23.3% of the drivers were between 50 -to-59 years old. This finding refuted a previous study which found that approximately 59% of bus drivers were aged between 50 and 59 years, with only 12.5% aged 60 years or above, and 28% under age 50 years [56]. This outcome implies that a significant proportion of the drivers are old and that complying with the road safety protocol would not be difficult for them to do [48]. The plausible reason for this outcome could probably be that these drivers might have been in the service since their youthful age and would want to retire in it. Further, it could also be that it has been their source of livelihood and that without it they would go to bed hungry [57].

It appeared Christian commercial drivers dominated in the study, hence, they constituted about sixty-two percent of the total sample. This finding refuted a previous study which found that 81.2% of the drivers identified as Christians, while 18.8% were Muslim [58]. This finding suggests that few of the drivers are affiliated to different religion [59]. The plausible explanation could be that the population of the study setting is dominated by Christianity [60]. However, the 11.1% that intimated they are traditionalist reason could be that they do not believe in the teachings of Jesus Christ [61]. This finding implies that some of the drivers do and perceive things in the traditional way [62]. This outcome is almost similar to a previous study which also found that 10.3% of their sample were believers in the African Traditional Religion [63].

Approximately one-third of the sampled individuals lived in households with five or more members. Consistent with previous studies, which found that in many regions—particularly sub-Saharan Africa, West Asia, South Asia, and Melanesia—households frequently have five or more members, and in some countries, the average exceeds six [64,65]. This outcome implies that a significant proportion of the drivers have a large number of family members they cater for [66–68]. This alone might make them sober to comply with the road safety protocol. Hence, knowing that you have a larger family depending on you for survival, you might not want to become a casualty to live them in distressed [69]. The plausible explanation to this finding could probably be that these drivers might be living with their extended families with their immediate family which make them to cater for other extended children in addition to their own children [68]. Further, it could be that both the man and the woman had children before they got married [70–72].

However, the 16.5% that indicated they belong to 2-member household reason could be that they just got married and that have not started making babies yet [73]. The plausible reason for this finding could be that these drivers want to be well established before they start to make babies [74]. This outcome refuted a previous study, which found that the breakdown of household sizes includes: 11.0% lived alone (single-member); 47.2% lived in 2–5member households.

The study found that more than half of the drivers have been in operation for more than six years. Consistent with previous studies, which found that the largest group (37.8%) had between 11 and 20 years of experience, indicating that the majority had well over six years behind the wheel [75].

This suggests that a significant proportion of the drivers are old in the transport industry, and that could commemorate with the experiences they have acquired to safeguard passengers’ safety in their daily journeys [76]. It could also imply why they comply with road safety protocol. The possible reason for this finding could be that these participants cherish working in the transport industry therefore, right after their junior high school or senior high school they went to driving school to learn to become professional drivers to help boost the transport industry [77].

On the other hand, the 5.8% participants that intimated they have been in operation for less than a year reason could be that they just completed their driving school and got hired [78]. Further, it could also be that they were working hard to get their own vehicle to start business hence, they do not want to work for anybody [79]. This finding implies that a significant proportion of the drivers are new in the transport industry and that need to be more careful to ensure that they can break all odds in order to be effective in the industry [80].

It emerged that 33% of the participants indicated that human factors such as physical and mental conditions can instigate road safety compliance among commercial drivers. This finding refuted a previous study, which found that approximately 25.5% of respondents admitted to driving while feeling sleepy [81]. The authors further stressed that sleep disorders such as obstructive sleep apnea (prevalence between 15–45% among drivers) and fatigue raise accident risks comparable to those caused by alcohol impairment. This outcome suggests that in the transport industry, one has to be physically healthy and mentally sound in order to be effective in the industry [82]. It could also suggest that to be able to adhere to road safety measures, one needs to be mentally sound to understand the need of complying with the safety protocol in the transport industry [83]. The plausible explanation to this finding could probably be that these drivers understand that most of the road traffic accidents that normally occur has to do with physical challenge/impairment and mentally derailed issues on the part of the victims [16,84].

However, the 17.0% that indicated drivers’ skills and experience reason could probably be that for one to be able to comply with the road safety protocol, one has to acquire some skills and experiences in order to be able to understand the road safety protocol before thinking of adhering to them [16,85]. This finding suggests that without skills and experience, one might not be able to comply with the road safety protocol, hence, they are complex to understand [86,87]. This finding is consistent with previous studies, which found that greater experience and higher formal education were linked to fewer violations, especially related to speeding or prohibited overtaking [88,89]. The authors further asserted that overall driving experience and type of training significantly influence rule adherence.

The research found that 39% of participants reported that vehicle conditions can trigger commercial drivers to comply with road safety protocol. Consistent with a previous study, which found that better-maintained vehicles encourage more cautious driving practices and compliance with safety rules [15]. The implication of this finding is that commercial drivers who comply with road safety protocol are mostly those who have their vehicles not in good shape [90–92]. The plausible explanation to this finding could be that those drivers might not want to get involved in a motor vehicle crash. So that in the end, authorities will ask them to remove their vehicles from the road hence, they are deteriorated or have outlived their life-span [93].

Road conditions play a major role in road safety protocol compliance. In view of this, about 45% of the participants endorsed that road conditions can influence them to comply with road safety protocol. Consistent with previous studies, which found that physical features perceived as restrictive (e.g., narrower lanes, high traffic, shoulder absence) significantly increased compliance—drivers adapted behaviour when road conditions signaled higher risk [94,95]. The authors further stressed that drivers were significantly more likely to stop or yield where features like speed humps, intersection lighting, and channelisation were present. This finding suggests that most of the roads that they use are not in good conditions and does not enhance easy plying [96]. The plausible explanation for this finding could probably be that the nature of the roads does humbles commercial drivers to comply with the road safety protocol [96,97].

On the other hand, the 22.3% that indicated lighting conditions reason could probably be that drivers who have issues with their headlights and other indicators do comply with the road safety protocol. Hence, they do not want to be labelled as victims of circumstance [98,99]. The implication of this finding is that a significant proportion of drivers have issues with their vehicle lights [100]. This underscores an urgent intervention to educate drivers on the need of why they must adhere to vehicle maintenance practices daily before the start of duty in every day [49,101,102]. This finding agrees with a previous study, which found that the presence of sufficient illuminated road signs significantly reduced perceived risk on roads outside the city by 22.3% (p < 0.05) and within the city by 14.6% (p < 0.01) [103].

The study found that about 39% of the participants reported that traffic volume could trigger them to adhere to road safety protocol. Consistent with a previous study, which found that high traffic volume was found to significantly increase compliance rates on both arterial and collector roads [94]. The authors further postulated that drivers were more likely to follow posted speed limits during peak traffic periods. This outcome implies that most of the roads they use have narrow lanes, have sharp curves, and do not also have sufficient signage [104–106]. The possible reason for this finding could probably be that these drivers normally encounter traffic jam anytime they are using the road which does not enhance the transport industry [107].

## Conclusion

The study brought to the fore that Islamic religion had weak association with road safety protocol compliance among commercial drivers at Hohoe in the Volta Region of Ghana. However, other variables were found to have played more influential roles. Notably, strong associations were identified between drivers’ compliance with road safety protocols and a range of factors including socio-demographic characteristics (such as age and years of experience), human factors (like mental and physical condition), vehicle conditions, environmental influences (such as lighting and traffic volume), and road-related features (including signage and road quality). A major limitation of the study is that it employed a quantitative cross-sectional design which did not permit the study to establish cause and effect. Also, the data collected was self-reported and may be influenced by respondent bias since their responses were based on the experiences they can remember.

## Declaration

### Ethical Approval

Ethical clearance (with ID (UHAS-REC A. [018]) 21-22) to carry out the study was obtained from the University of Health and Allied Sciences, Ho, Ghana.

### Consent to participate in the Study

In the field, verbal consent was sought before a participant could take part in the study.

### Consent to Publish

Participants were informed that the study was strictly for academic purposes and that the results would be published for the purposes of contributing to building academic literature.

### Competing Interests

No competing interest existed.

### Funding

The study was self-funded

### Availability of Data and Materials

The data is only available to the authors hence it was a primary data. However, it can be shared upon request from the corresponding author through, University of Cape Coast, Cape Coast, Ghana.

## Data Availability

The data is primary and contains sensitive information about the participants which prohibits sharing publicly. However, it can be shared upon request from the corresponding author through

## Acknowledgements

Sincerely, we are grateful to the respondents who sacrifice their time to take part in the study and the research assistants for their help during the data collection.

## Authors’ Contribution

This was an Undergraduate Research by AAA who collected the data, did the analysis, and wrote the initial draft of the manuscript. VOC and AAA Conceptualize the study. VOC supervised the dissertation and assisted in the Methodology. AEB wrote the original draft of the manuscript from the dissertation for publication. FA, and AEB critically reviewed the manuscript to improve its scientific quality. All authors contributed to a revision of the manuscript and gave consent for its publication.

## Notes

### Competing Interest Statement

The authors have declared no competing interest.

### Funding Statement

The author(s) received no specific funding for this work.

### Author Declarations

The study obtained an ethical clearance with ID (UHAS-REC A. [018]) 21-22 from The Research Committee of the University of Health and Allied Sciences

